# Plasma proteomic signatures for type 2 diabetes mellitus and related traits in the UK Biobank cohort

**DOI:** 10.1101/2024.09.13.24313501

**Authors:** Trisha P. Gupte, Zahra Azizi, Pik Fang Kho, Jiayan Zhou, Kevin Nzenkue, Ming-Li Chen, Daniel J. Panyard, Rodrigo Guarischi-Sousa, Austin T. Hilliard, Disha Sharma, Kathleen Watson, Fahim Abbasi, Philip S. Tsao, Shoa L. Clarke, Themistocles L. Assimes

**Author notes:** **Corresponding author:** Themistocles L. Assimes, MD PhD Palo Alto VA Hospital, 3801 Miranda Avenue, Palo Alto, CA, 94304.

## Abstract

**Aims/hypothesis:** The plasma proteome holds promise as a diagnostic and prognostic tool that can accurately reflect complex human traits and disease processes. We assessed the ability of plasma proteins to predict type 2 diabetes mellitus (T2DM) and related traits.

**Methods:** Clinical, genetic, and high-throughput proteomic data from three subcohorts of UK Biobank participants were analyzed for association with dual-energy x-ray absorptiometry (DXA) derived truncal fat (in the adiposity subcohort), estimated maximum oxygen consumption (VO_2_max) (in the fitness subcohort), and incident T2DM (in the T2DM subcohort). We used least absolute shrinkage and selection operator (LASSO) regression to assess the relative ability of non-proteomic and proteomic variables to associate with each trait by comparing variance explained (R^2^) and area under the curve (AUC) statistics between data types. Stability selection with randomized LASSO regression identified the most robustly associated proteins for each trait. The benefit of proteomic signatures (PSs) over QDiabetes, a T2DM clinical risk score, was evaluated through the derivation of delta (Δ) AUC values. We also assessed the incremental gain in model performance metrics using proteomic datasets with varying numbers of proteins. A series of two-sample Mendelian randomization (MR) analyses were conducted to identify potentially causal proteins for adiposity, fitness, and T2DM.

**Results:** Across all three subcohorts, the mean age was 56.7 years and 54.9% were female. In the T2DM subcohort, 5.8% developed incident T2DM over a median follow-up of 7.6 years. LASSO-derived PSs increased the R^2^ of truncal fat and VO_2_max over clinical and genetic factors by 0.074 and 0.057, respectively. We observed a similar improvement in T2DM prediction over the QDiabetes score [Δ AUC: 0.016 (95% CI 0.008, 0.024)] when using a robust PS derived strictly from the T2DM outcome versus a model further augmented with non-overlapping proteins associated with adiposity and fitness. A small number of proteins (29 for truncal adiposity, 18 for VO2max, and 26 for T2DM) identified by stability selection algorithms offered most of the improvement in prediction of each outcome. Filtered and clustered versions of the full proteomic dataset supplied by the UK Biobank (ranging between 600-1,500 proteins) performed comparably to the full dataset for T2DM prediction. Using MR, we identified 4 proteins as potentially causal for adiposity, 1 as potentially causal for fitness, and 4 as potentially causal for T2DM.

**Conclusions/Interpretation:** Plasma PSs modestly improve the prediction of incident T2DM over that possible with clinical and genetic factors. Further studies are warranted to better elucidate the clinical utility of these signatures in predicting the risk of T2DM over the standard practice of using the QDiabetes score. Candidate causally associated proteins identified through MR deserve further study as potential novel therapeutic targets for T2DM.

## Introduction

As its incidence rises, a critical need exists to improve our ability to risk stratify and prevent type 2 diabetes mellitus (T2DM) [1]. Insulin resistance (IR) is characterized as a decreased sensitivity to insulin-mediated glucose uptake and is a known primary risk factor for T2DM [2, 3]. While more reliable identification of those with IR could prove useful for T2DM risk stratification, direct measures of IR remain both expensive and laborious to perform [4, 5] and surrogate measures correlate only modestly with direct measures of IR [6–8]. Truncal adiposity and poor cardiorespiratory fitness (CRF) are two additional potentially modifiable risk factors of T2DM through their effects on IR but similar to IR, are difficult to accurately measure using gold standard approaches such as dual energy x-ray absorptiometry (DXA) scans and cardiopulmonary exercise testing with a metabolic cart [9–12].

In the last decade, high-throughput profiling of circulating plasma proteins has emerged as a powerful tool for both predicting and understanding the underlying biology of complex traits and incident disease [13]. The incorporation of plasma proteomic data to routinely available clinical information has also been shown to improve prediction of cardiometabolic traits in particular [14–16]. However, the ability of proteins to enhance prediction beyond that offered by more robust T2DM clinical risk prediction models such as the UK-based QDiabetes clinical risk score (which incorporates significant past medical history, past medication use, and other clinical variables in individuals) has yet to be tested [17].

Recently, our laboratory established plasma proteomic signatures of a direct measure of IR, which were obtained in two smaller-sized population cohorts [18]. We sought to expand on this work by leveraging clinical, genetic, and proteomic data within a substantially larger cohort, the UK Biobank (UKB). As the UKB currently lacks any direct measures of IR, we assessed the relative ability of proteins to predict two risk factors upstream of IR, truncal adiposity and CRF, as well as a downstream consequence of IR, T2DM. In predicting T2DM, we hypothesized that the inclusion of proteomic data would provide modest enhancement in prediction accuracy beyond that offered by the QDiabetes risk score and other traditional clinical and genetic data.

## Methods

### Study design and population

The study design of the UK Biobank (UKB) has been previously described extensively [19]. At participants’ baseline visits, trained healthcare providers conducted verbal interviews and administered questionnaires to obtain information on health status, family history, sociodemographic and psychosocial factors, and lifestyle. Physical measures along with the collection of blood, urine, and saliva were also obtained at these visits. By integrating participants’ electronic health records (EHRs), health outcomes data including outpatient and inpatient International Classification of Disease, Tenth Revision (ICD-10) codes are also available within the database. The UKB received ethical approval from the Northwest Multicenter Research Ethics Committee and obtained informed consent from all participants at the time of recruitment.

We analyzed three non-overlapping subcohorts of UKB participants who underwent high-throughput proteomic profiling to study outcomes of truncal adiposity, fitness, and T2DM (**ESM Fig. 1**). First, individuals who underwent a DXA scan at the imaging visit comprised an adiposity subcohort (n = 5,506). Next, individuals who completed cycle ergometer tests at the baseline visit comprised a fitness subcohort (n = 5,559). Finally, the remaining individuals who had proteomics data but did not undergo DXA scans or cycle ergometer tests comprised a T2DM subcohort (n=35,946). Individuals with prevalent T2DM, any history of prevalent or incident type I diabetes mellitus (T1DM), or missing or elevated A1c (≥48 mmol/mol [6.5%]) values were excluded from this study. Individuals in whom the QDiabetes score could not be computed were also excluded from the T2DM subcohort.

**Figure 1.**
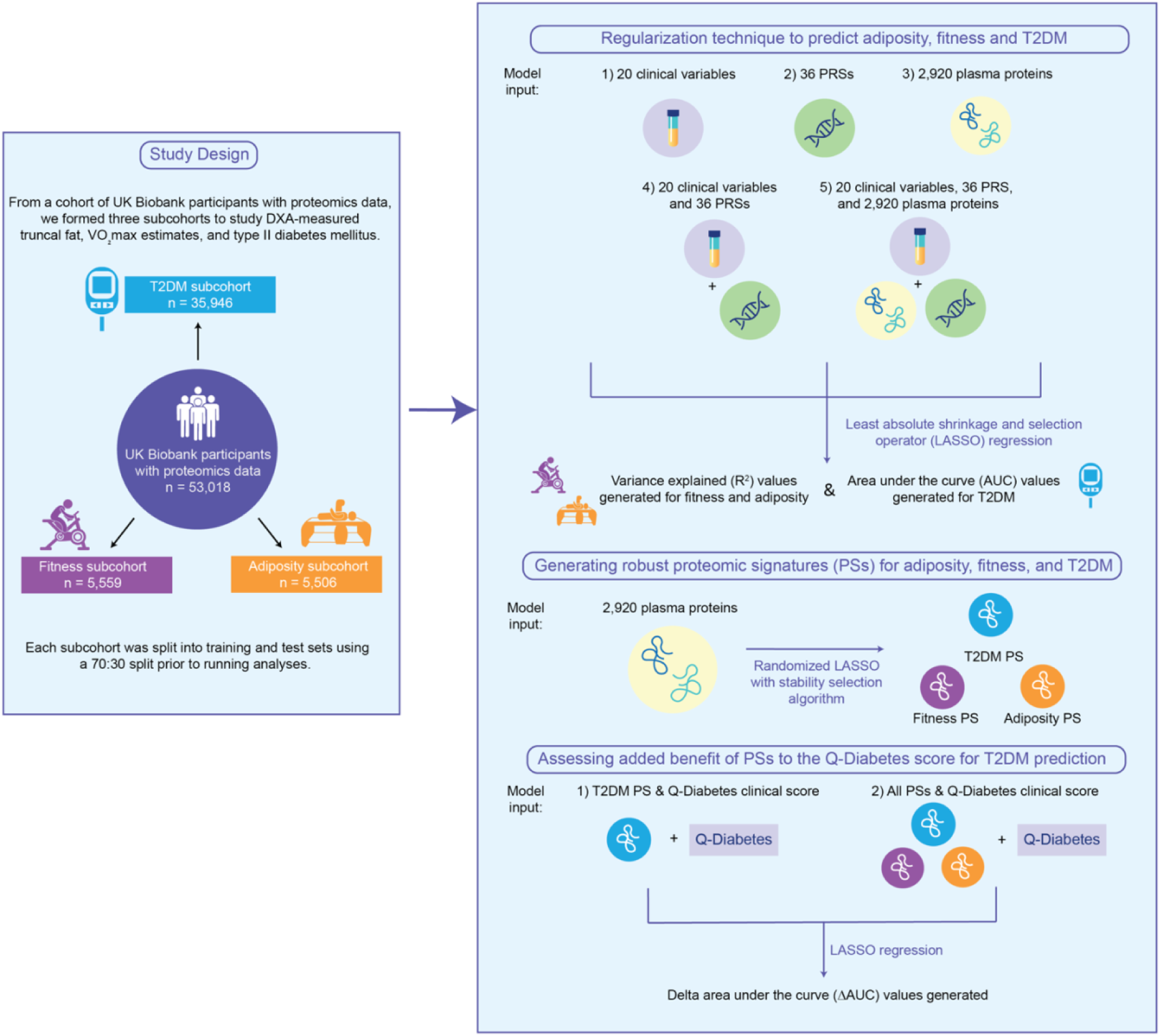
Study design and analysis workflow. **Abbreviations:** DXA: dual-energy x-ray absorptiometry, T2DM: type 2 diabetes mellitus, T1DM: type I diabetes mellitus, A1c: hemoglobin A1c, VO_2_max: maximal oxygen consumption

### Measurement of protein biomarkers

Using the antibody-based Proximity Extension Assay by Olink, the UKB measured normalized protein expression (NPX) data for a total of 2,923 circulating proteins in plasma in a subset of UKB participants. The sample handling, processing, and quality control protocols implemented by the UKB have been previously described in a summary document (biobank.ndph.ox.ac.uk/ukb/ukb/docs/PPP_Phase_1_QC_dataset_companion_doc.pdf) and in two publications [20, 21]. We identified GLIPR1, NPM1, and PCOLCE as proteins with a high degree of missingness (> 50%) and excluded these from analysis. The remaining missing NPX values were imputed with their mean values. All NPX values were provided as log-transformed values by the UKB and then standardized by us prior to analysis.

### Measurement of outcomes

Adiposity was defined as DXA scan-measured truncal fat tissue percentage (equal to trunk fat mass divided by total fat mass). Fitness was defined as an estimate of maximal oxygen consumption (VO_2_max). These estimates were previously generated and validated by Gonzales et al. using a multilevel modelling framework that incorporated participants’ heart rate responses during the fitness test [22]. More details on the DXA scan and fitness test protocols employed by the UKB can be found in the electronic supplementary material (**ESM Methods**). Incident T2DM was defined as any instance of an E11 ICD-10 code following participants’ baseline visits within UKB’s first occurrences data (UKB categories 2401-2417). This data is organized by ICD-10 codes and was generated by mapping primary care data (UKB category 3000), hospital inpatient data (UKB category 2000), death register records (UKB fields 40001 and 40002), and self-reported medical condition codes (UKB field 20002).

### Measurement of clinical variables

We selected clinical variables to use as predictors in the analysis after reviewing known predictors of each trait within the literature. The following groups of clinical variables documented at baseline visits were included in our analyses: demographic characteristics (sex, age, self-reported ethnicity, and Townsend deprivation scores (TDSs)), past medical history, family history, medication use, health behaviors (prior alcohol use, ever smoked status, and physical activity levels), and physical measures (weight, body mass index (BMI), waist circumference, and systolic and diastolic blood pressure measurements). From blood and saliva samples obtained during baseline visits, we also included standard biochemistry markers, and 36 standard polygenic risk scores (PRSs) generated from external genome-wide association studies [23, 24]. In the T2DM subcohort, QDiabetes, a clinical risk prediction model previously validated in UK-based populations for T2DM prediction, was also included as a clinical variable [17]. We computed this score using the QDiabetes 2018C function within the QDiabetes R package. We standardized all clinical variables and PRSs prior to running analyses. A full list of clinical variables and PRSs used in each of the subcohorts are included in **ESM Methods**.

### Statistical analyses

A flowchart of the study design and analysis plan is shown in **Figure 1**. We used least absolute shrinkage and selection operator (LASSO) regression to assess the relative ability of clinical variables, PRSs, and proteins to associate with adiposity, fitness, and T2DM. After randomly splitting each subcohort into training (70%) and test (30%) sets, all LASSO models were built using 10-fold cross validation. In each subcohort, we developed five LASSO models: a model with clinical variables alone, a model with PRSs alone, a model with proteins alone, a combined model in which PRSs were added to clinical variables, and a combined model in which proteins were added to clinical variables and PRSs. In the adiposity and fitness subcohorts, model improvement was defined as an increase in variance explained (R^2^) and in the T2DM subcohort, as an increase in the area under the curve (AUC) value. To assess the added incremental value of proteins beyond routinely available clinical variables and PRSs, we derived a delta (Δ) AUC value and generated a corresponding 95% confidence interval by bootstrapping 1,000 samples.

Next, we implemented the randomized LASSO stability selection (RLSS) algorithm using the R package stabs, which was initially presented by Meinhausen and Bühlmann and later improved upon by Shah and Samsworth [25, 26]. This algorithm was applied in training sets to generate a more robustly associated proteomic signature (PS) for each trait of interest. We used default parameters when applying this algorithm, which included a weakness value of 0.8, a cutoff value of 0.8, and a per-family error rate of two. To determine how much prediction was offered by these smaller sets of proteins, we measured the R^2^ of LASSO models run with the adiposity and fitness-associated PSs and the AUC of a LASSO model run with the T2DM PS in the test sets of each respective subcohort. Next, we again derived Δ AUC values with 95% confidence intervals to determine the added benefit of these PSs in prediction of T2DM beyond that offered by the QDiabetes clinical risk score.

To further assess correlation structure and the incremental gain in model performance with proteomic datasets of varying numbers of included proteins, we used two methods to reduce high protein-protein correlation within the full proteomic dataset provided by UKB. In a filtering-based approach, we first computed a correlation matrix of all 2,920 plasma proteins and identified pairs of proteins with a correlation value > 0.3 and > 0.5. Next, we randomly removed one protein from each of these pairs to form two smaller datasets of approximately 600 and 1,500 proteins. In the second clustering-based approach, we used principal component analysis (PCA) and K-means clustering to form 600 and 1,500 clusters of proteins. Next, we randomly selected one protein from each cluster to form two additional smaller datasets of 600 and 1,500 proteins. Finally, using standard LASSO regression, we compared T2DM prediction performance of all four proteomic datasets by generating AUCs.

We conducted a series of two-sample Mendelian randomization (MR) analyses to identify potentially causal plasma proteins for adiposity, fitness, and T2DM (**ESM Fig. 2**). We first performed genome-wide association studies (GWASs) for all 2,920 proteins in an independent cohort of 15,016 UKB participants to identify *cis-*protein quantitative trait loci (*cis*-pQTLs).

**Figures 2a-b.**
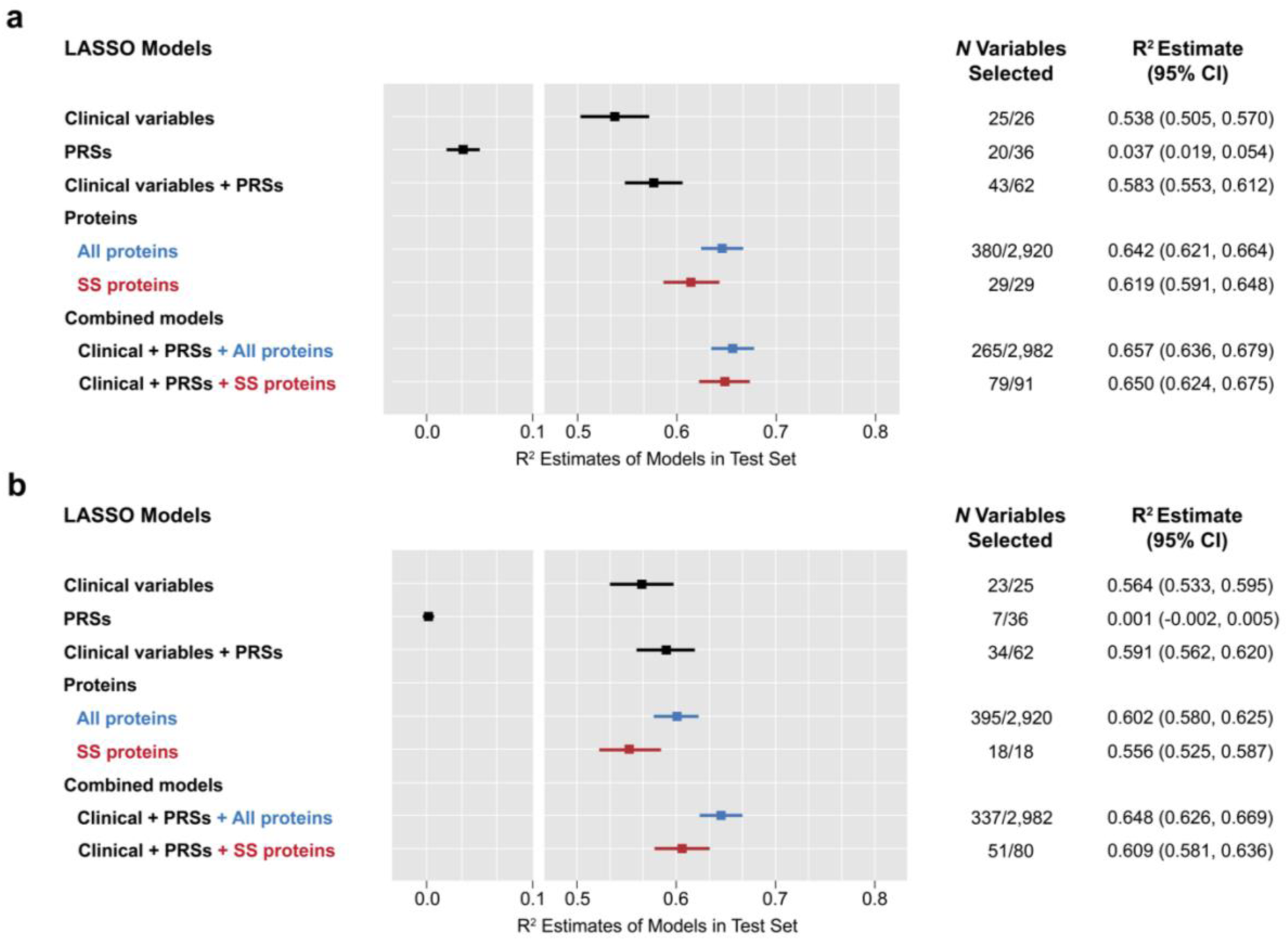
Variance explained (R^2^) of clinical variables, polygenic scores, and plasma proteins in the adiposity and fitness subcohorts. **Footnote:** (**a**) Models performed using training and test sets in the adiposity subcohort. (**b**) Models performed using training and test sets in the fitness subcohort. SS proteins shown in red refer to proteins selected by a randomized LASSO regression model with stability selection algorithm. **Abbreviations:** LASSO: least absolute shrinkage and selection operator, R^2^: variance explained, PRSs: polygenic risk scores, SS: stability selection

Effect estimates for truncal adiposity and VO_2_max were obtained by performing GWAS analyses of DXA-measured truncal fat percentage and VO_2_max estimates in a cohort of 33,348 UKB participants and a cohort of 62,402 UKB participants, respectively. Finally, effect estimates for T2DM were obtained from a meta-analysis of summary statistics published by the DIAMANTE (DIAbetes Meta-ANalysis of Trans-Ethnic association studies) consortium (n_cases_ = 55,005 & n_controls_ = 400,308) [27]. Details on the cohorts and methods used for all three GWAS analyses and on the DIAMANTE consortium’s meta-analyzed summary statistics can be found in **ESM Methods**.

We used the inverse variance weighted (IVW) method as our primary method to conduct the series of two-sample MR analyses. There are three core assumptions which should be met in a MR analysis: 1) the genetic variants used as instrumental variables should be strongly associated with the outcome of interest, 2) the genetic instruments should not be associated with other confounder variables, and 3) the genetic instruments should impact the outcome of interest only via the exposure of interest. While MR analyses using *cis*-genetic variants are generally regarded to be assumption satisfied, we addressed these assumptions through additional analyses [28].

First, we assessed the strength of our genetic instruments by calculating the proportion of variance explained and *F* statistics using previously established methods. The equations used to calculate both can be found in **ESM Methods.** We also ran additional sensitivity analyses, including the MR-Egger method and specifically, calculating the MR-Egger intercept to test for pleiotropy. All analyses were conducted using the TwoSampleMR package in R.

## Results

We analyzed NPX data of 2,920 proteins in a total of 47,011 participants. Baseline characteristics for each subcohort are shown in **Table 1**. Across all three subcohorts, the mean age at recruitment was 56.65 years (SD, 8.22 years), 54.9% were female, and 94.2% were of European ancestry. In the T2DM subcohort, 5.8% developed incident T2DM over a median follow-up of 7.6 years.

**Table 1.** Demographics and clinical characteristics of the study population in each subcohort.

| **n** | **Adiposity**<br>**subcohort**<br>**(n = 5,506)** | **Fitness**<br>**subcohort**<br>**(n = 5,559)** | **T2DM**<br>**subcohort**<br>**(n = 35,946)** |
| --- | --- | --- | --- |
| Female (%) | 2,936 (53.3) | 2,958 (53.2) | 19,905 (55.4) |
| Age at baseline visit | 54.3 (7.9) | 56.8 (8.2) | 57.0 (8.2) |
| Self-reported ethnicity group (%) |  |  |  |
| White | 5,320 (96.6) | 5,034 (90.6) | 33,910 (94.4) |
| African | 28 (0.5) | 91 (1.6) | 369 (1.0) |
| South Asian | 45 (0.8) | 109 (2.0) | 437 (1.2) |
| Chinese | 11 (0.2) | 25 (0.4) | 99 (0.3) |
| Mixed | 38 (0.7) | 61 (1.1) | 205 (0.6) |
| Other | 64 (1.2) | 235 (4.2) | 902 (2.5) |
| BMI (kg/m<sup>2</sup>) | 26.4 (4.1) | 27.0 (4.3) | 27.4 (4.7) |
| Waist circumference (cm) | 87.4 (12.2) | 89.3 (12.7) | 90.1 (13.1) |
| SBP (mmHg) | 134.0 (16.9) | 136.5 (17.3) | 138.1 (18.2) |
| DBP (mmHg) | 81.0 (9.6) | 81.7 (9.7) | 82.4 (9.8) |
| LDL (mmol/mol) | 3.6 (0.8) | 3.6 (0.8) | 3.6 (0.9) |
| HDL (mmol/mol) | 1.8 (0.4) | 1.5 (0.4) | 1.4 (0.4) |
| TG (mmol/mol, median [IQR]) | 1.4 [0.4, 2.3] | 1.4 [0.4, 2.4] | 1.5 [0.4, 2.6] |
| Cholesterol (mmol/mol) | 4.7 (0.9) | 4.7 (0.9) | 4.7 (1.0) |
| HbA1c (mmol/mol) | 34.4 (3.5) | 35.1 (3.8) | 35.3 (3.9) |
| Blood pressure lowering medication (%) | 414 (7.5) | 572 (10.3) | 3,966 (11.0) |
| Cholesterol lowering medication (%) | 388 (7.0) | 566 (10.2) | 3,494 (9.7) |
| Past medical history of HTN (%) | 970 (17.6) | 1,347 (24.3) | 9,725 (27.1) |
| Past medical history of CVD (%) | 106 (1.9) | 184 (3.3) | 1,434 (4.0) |
| Family history of T2DM (%) | 996 (18.1) | 1,172 (21.1) | 7,264 (20.2) |
| Ever smoked status (%) | 3,218 (58.4) | 3,291 (59.2) | 21,569 (60.0) |
| Self-reported alcohol intake (%) |  |  |  |
| Never or missing | 265 (4.8) | 454 (8.2) | 3,136 (8.7) |
| One to three times a month or special occasions | 1,007 (18.3) | 1,231 (22.1) | 8,129 (22.6) |
| Once or twice a week | 1,425 (25.9) | 1,393 (25.1) | 9,479 (26.4) |
| Three or four times a week | 1,546 (28.1) | 1,329 (23.9) | 7,906 (22.0) |
| Daily or almost daily | 1,263 (22.9) | 1,152 (20.7) | 7,296 (20.4) |
| Physical activity category (%) |  |  |  |
| Missing | 850 (18.4) | 1,169 (21.0) | 8,576 (23.9) |
| Low | 855 (15.5) | 671 (15.3) | 5,144 (18.8) |
| Moderate | 2463 (44.7) | 2,298 (52.3) | 13,616 (49.7) |
| High | 1338 (24.3) | 1,421 (32.4) | 8,610 (31.5) |
| Total truncal fat tissue percentage | 37.2% (9.6%) | - | - |
| VO<sub>2</sub>max estimate (mL/kg/min) |  | 28.4 (6.8) |  |
| Incident T2DM | - | - | 2,077 (5.8) |
| Follow-up time in years | - | - | 7.6 (3.9) |

All continuous measurements were documented in mean (SD) unless otherwise specified.
**Abbreviations:** IQR: interquartile range, BMI: body mass index, SBP: systolic blood pressure, DBP: diastolic blood pressure, LDL: low-density lipoprotein, HDL: high-density lipoprotein, TG: triglyceride, HbA1c: hemoglobin A1c, HTN: hypertension, CVD: cardiovascular disease, T2DM: type 2 diabetes mellitus, VO<sub>2</sub>max: maximal oxygen consumption

### Truncal adiposity analyses

Standard LASSO models built on clinical variables alone, PRSs alone, or the full proteomic dataset on its own explained 0.538, 0.037, and 0.642 of variance in adiposity, respectively (**Fig. 2a**). A LASSO model combining both clinical variables and PRSs explained 0.583 of variance (R^2^) in adiposity. Thus, we found that a model incorporating the full proteomic dataset on its own performed notably better than a model built on clinical variables and PRSs. Incorporating the full proteomic dataset in addition to routinely available clinical variables and PRSs increased the R^2^ value from 0.583 to 0.657. The randomized LASSO stability selection (RLSS) analysis selected 29 proteins (**Fig. 3a**), and these proteins offered most of the improvement in prediction of adiposity (R^2^=0.623). A full list of the clinical variables, PRSs, and proteins selected by these LASSO models can be found in **ESM Table 1**.

**Figure 3a-c.**
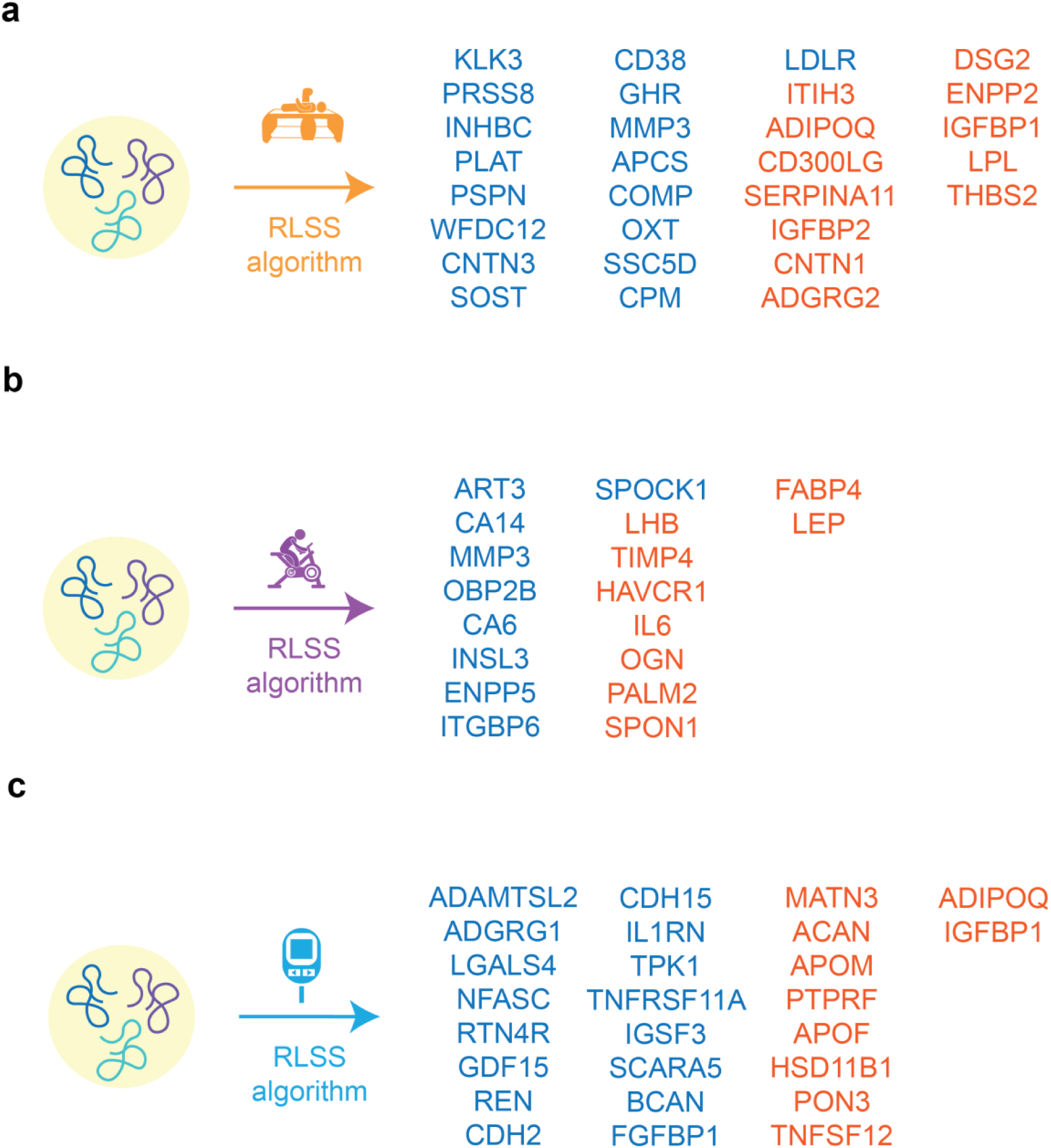
Proteins selected by a randomized LASSO with stability selection algorithm for adiposity, fitness, and type 2 diabetes mellitus. **Footnote:** (**a**) Proteins selected by RLSS algorithm in the adiposity subcohort. (**b**) Proteins selected by RLSS algorithm in the fitness subcohort. (c) Proteins selected by RLSS algorithm in the T2DM subcohort. Proteins listed in blue were positively associated with the outcome of interest while those listed in orange were negatively associated. **Abbreviations:** RLSS: Randomized LASSO regression with stability selection algorithm.

### Cardiorespiratory fitness analyses

Standard LASSO models built on clinical variables alone, PRSs alone, or the full proteomic dataset on its own explained 0.564, 0.001, and 0.602 of variance in estimated VO_2_max, respectively (**Fig. 2b**). A LASSO model incorporating both clinical variables and PRSs had an R^2^ value of 0.591. We found that a protein-only model performed similarly in prediction of fitness to a model combining clinical variables and PRSs. Incorporating proteins in addition to routinely available clinical variables and PRSs increased the R^2^ value from 0.591 to 0.648. RLSS analysis selected 18 proteins (**Fig. 3b**) and similar to results from our adiposity analyses, this substantially smaller set of proteins explained the majority of fitness variance with an R^2^ value of 0.556. A full list of the clinical variables, PRSs, and proteins selected by these LASSO models can be found in **ESM Table 2**.

### Type 2 diabetes mellitus analyses

Consistent with prior reports, the QDiabetes clinical risk score performed very well on its own in predicting T2DM with an AUC of 0.865 (95% CI 0.851-0.880) (**Fig. 4a**). When additional clinical variables were added to the QDiabetes score, T2DM prediction improved with an AUC of 0.872 (95% CI 0.858-0.886). In comparison to this clinical-variable only model, a PRS-only model performed much worse with an AUC of only 0.666 (95% CI 0.645-0.687) while a model built with the full proteomic dataset on its own performed slightly worse with an AUC of 0.859 (95% CI 0.845-0.873). When combining additional clinical variables not already in the QDiabetes score and PRSs with the QDiabetes score, we observed a modest increase in AUC to 0.876 (95% CI 0.864-0.891). A standard LASSO regression model incorporating proteins performed similarly to the QDiabetes score alone with an AUC of 0.859 (95% CI 0.845-0.873). Incorporating proteins in addition to the QDiabetes score, other routinely available clinical variables, and PRSs resulted in a Δ AUC of only 0.014 (95% CI 0.007-0.024).

**Figures 4a-d.**
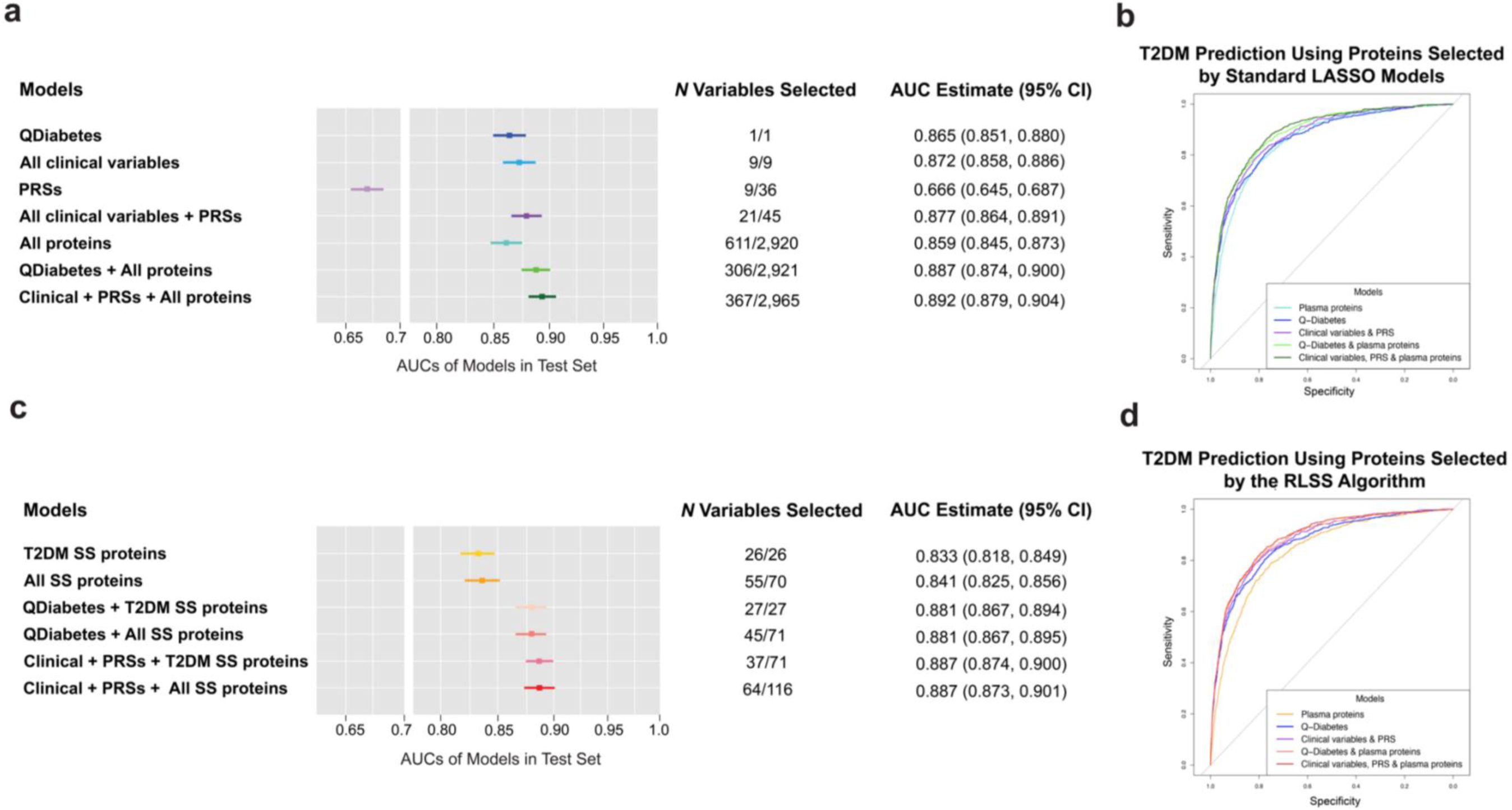
Area under the curves (AUCs) of clinical variables, polygenic scores, and plasma proteins in the type 2 diabetes mellitus subcohort. **Footnote:** (**a, b**) Models performed in training and test sets of the T2DM subcohort using the full proteomic dataset. (**c, d**) Models performed in training and test sets of the T2DM subcohort using proteins selected by a randomized LASSO regression model with a stability selection algorithm. Colors shown on the receiver operating curves correspond with colors shown on the forest plots. **Abbreviations:** LASSO: least absolute shrinkage and selection operator, T2DM: type 2 diabetes mellitus, AUC: area under the curve, PRS: polygenic risk scores, SS: stability selection

RLSS analysis selected 26 proteins (**Fig. 3c**), which offered most of the improvement in T2DM prediction explained by the full proteomic dataset with an AUC of 0.833 (95% CI 0.818-0.849) (**Fig. 4b**). When this proteomic signature was augmented with non-overlapping stability selection proteins associated with adiposity and fitness, T2DM prediction marginally improved with an AUC of 0.837 (95% CI 0.822-0.853). Incorporating these three proteomic signatures in addition to QDiabetes resulted in a Δ AUC of 0.016 (95% CI 0.008-0.024). A full list of clinical variables, PRSs, and proteins selected by these LASSO models can be found in **ESM Table 3**.

### Clustering and filtering analyses

We assessed and compared the prediction performance of four smaller subsets of proteins formed through filtering-based or clustering-based approaches to that of the full proteomic dataset provided by UKB [AUC of 0.865 (95% CI 0.851-0.880)] (**ESM Fig. 3**). A subset formed through a filtering-based approach with a correlation threshold of > 0.3 (600 proteins included) performed the worst with an AUC of 0.815 (95% CI 0.798-0.832). Despite consisting of approximately the same number of proteins (645 vs. 600), a subset formed through a clustering-based approach with the creation of 600 clusters performed better with an AUC of 0.845 (95% CI 0.830-0.861). Finally, subsets formed through a filtering-based approach with a correlation threshold of > 0.5 and through a clustering-based approach with the creation of 1,500 clusters performed comparably to the full proteomic dataset with AUCs of 0.851 (95% CI 0.798-0.832) and 0.845 (95% CI 0.830-0.861), respectively.

### Two-sample Mendelian randomization analyses

To identify potentially causal proteins for adiposity, fitness, and T2DM, we conducted a series of two-sample Mendelian randomization (MR) analyses. Of 2,920 proteins, we identified genome-wide significant *cis*-protein quantitative loci (*cis*-pQTLs) for 1,745 based on a significance threshold of 5 x 10^-8^. The minimal *F* statistic of instrumental variables used in each analysis was 20.6 (**ESM Tables 4-6**). Effect estimates for adiposity and fitness were obtained by running GWAS analyses using the REGENIE software (**ESM Fig. 4a-d**) while effect estimates for T2DM were obtained from meta-analyzed summary statistics provided by the DIAMANTE consortium. We identified 3 proteins as potentially causal for truncal adiposity, one protein as potentially causal for fitness, and 4 proteins as potentially causal for T2DM based on FDR < 0.05 (**Fig. 5a-c**). For several proteins in which the initial number of associated SNPs was low (nSNPs < 3), we were not able to obtain results for sensitivity analyses. For proteins with a higher number of initial associated SNPs, however, we found that results from the IVW method generally aligned with results from other sensitivity analyses (**ESM Fig. 8-10**). Full results for each two-sample MR analysis including results from sensitivity analyses and annotations of whether each protein tested was selected by a standard LASSO model or RLSS analysis can be found in the supplementary materials (**ESM Tables 4-6**).

**Figure 5a-c.**
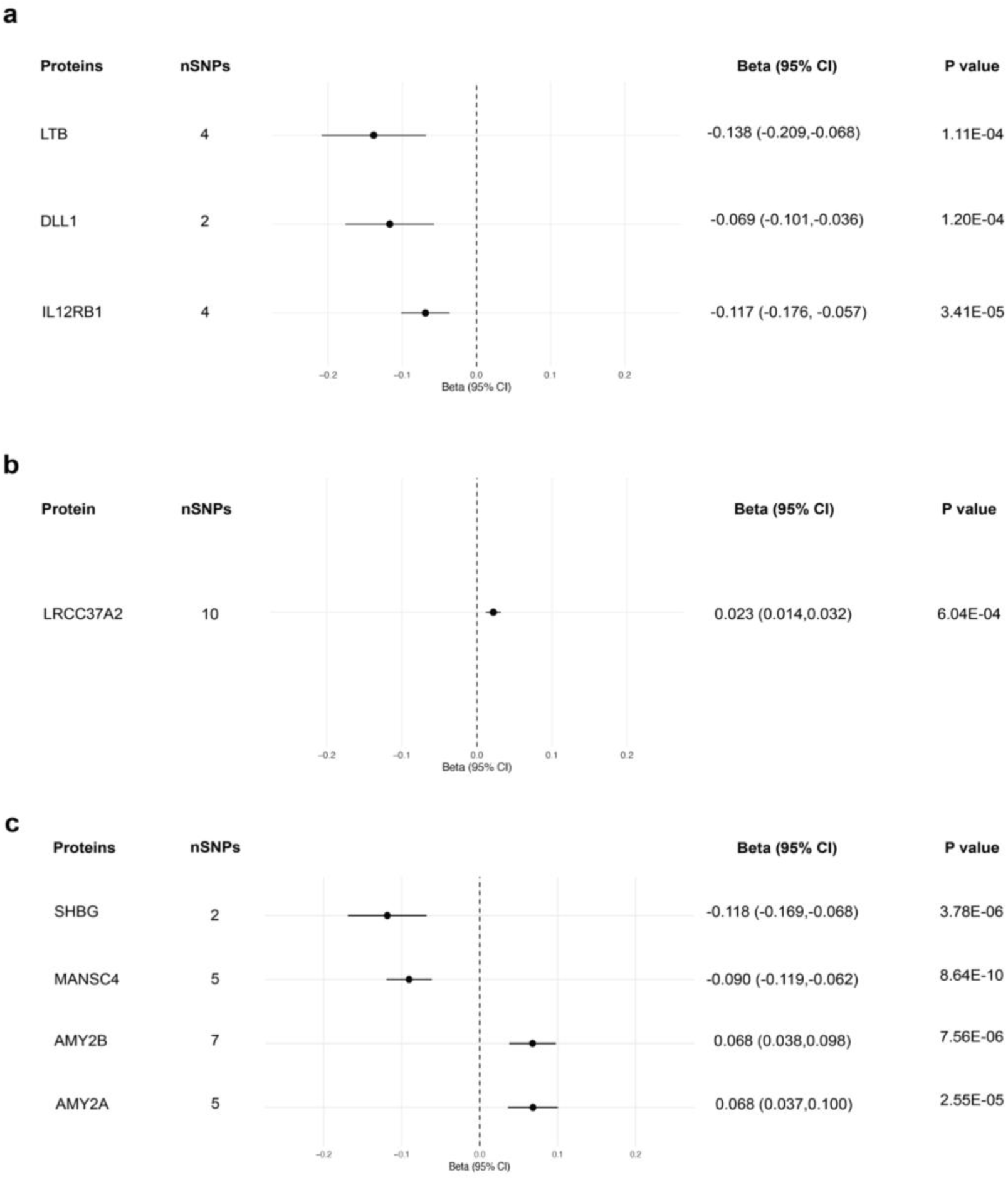
Potentially causal proteins for adiposity, fitness, and type 2 diabetes mellitus identified through two-sample Mendelian randomization. **Footnote:** (a) Forest plot of potentially causal proteins for truncal adiposity based on FDR < 0.5. (b) Forest plot of a potentially causal protein for cardiorespiratory fitness based on FDR < 0.5. (c) Forest plot of potentially causal proteins for T2DM based on FDR < 0.5. **Abbreviations:** SNP: single nucleotide polymorphism, 95% CI: 95% confidence interval, T2DM: type 2 diabetes mellitus

## Discussion

We aimed to develop and assess the prediction performance of proteomic signatures for two risk factors upstream of IR, truncal adiposity and CRF, as well as a downstream consequence of IR, T2DM, using high-throughput proteomic profiling data provided by the UKB. In doing so, we tested whether proteomic data would enhance prediction accuracy of all three traits beyond that offered by an existing clinical risk prediction model, as well as traditional clinical and genetic factors. From our results, we highlight three principal sets of findings.

First, protein-only models derived by standard LASSO regression explained greater variance in both truncal adiposity and CRF compared to models restricted to clinical variables and PRSs. Models that combined proteins with clinical variables and PRSs also improved the R^2^ of both traits beyond that offered by models restricted to only clinical variables and PRSs. These findings suggest that proteins add incremental benefit to clinical variables and PRSs for prediction of both traits. Further, in settings in which routine clinical information may not be available, proteomic signatures similar to those we identified for truncal adiposity and CRF could serve as surrogate measures for both traits.

Second, we found that a protein-only model derived by standard LASSO regression performed similarly for incident T2DM prediction to the QDiabetes score which notably, performed exceedingly well on its own. A model that combined other clinical variables and PRSs with the QDiabetes score modestly enhanced T2DM prediction, while the subsequent addition of proteins resulted in a further modest improvement. Unlike most other T2DM risk scores, the UK-based QDiabetes score incorporates a wealth of data on past medical conditions and even current use of medications known to increase T2DM risk. As such, the score has previously demonstrated excellent performance in UK-based populations such as its validation cohort and in the UKB [17, 29]. Given the already strong performance of the QDiabetes score on its own, our observation of only modest improvements in T2DM prediction with the addition of other clinical variables and multi-omic datasets is not unexpected.

With this set of findings, however, we note a few additional considerations. First, others have previously demonstrated that metrics such as the AUC are often too under-powered to accurately quantify improvement in prediction model performance with the addition of novel biomarkers [29]. These limitations are further exacerbated when the baseline set of risk predictors already demonstrates exceptional performance. Thus, in the context of predicting a highly prevalent condition such as T2DM, it is certainly plausible that the marginal increases we demonstrate in AUC for T2DM prediction may not fully reflect the advantages offered by multi-omic datasets. Ultimately, other large-scale studies and more ideally, a prospective cohort study may be needed to best assess this benefit. Lastly, when looking to health systems that lack a cohesive integration of EHR databases and scenarios in which practitioners do not have access to the significant amount of clinical data needed to compute the QDiabetes risk score, the comparative performance of our protein-only model suggests that proteins may serve as a suitable alternative in the future.

Third, we demonstrate that smaller sets of proteins perform comparably to the full proteomic dataset provided by the UKB for prediction of all three traits. Across all three traits, substantially smaller sets of proteins selected by a stability selection algorithm accounted for most of the prediction performance offered by the full dataset. Aside from aiding in feature reduction, this method yields additional benefits by selecting more “stable” proteins which have a greater likelihood of being generalizable to other populations. Further, a future where robust proteomic signatures similar to those we describe may be selected for absolute quantification is foreseeable given the recent advent of custom quantitative PEA panels such as the CVD-21 tool [30]. We also show that using smaller datasets created by filtering and clustering perform comparably to the full dataset for T2DM prediction. Taken together, our findings suggest that the significant costs and time associated with measuring nearly 3,000 proteins may be avoided in future implementations of plasma proteomic profiling without meaningfully affecting prediction performance.

Recently, others have shown that employing a cross-trait integration of PRSs can improve risk prediction of a given target trait [31]. Indeed, we found that the inclusion of 36 PRSs spanning a wide range of conditions led to a modest improvement in T2DM prediction. Moreover, for prediction of each trait, we observed a pattern of PRSs for autoimmune conditions such as rheumatoid arthritis, celiac disease, and systemic lupus erythematosus along with various cancers often being selected by LASSO. The role of pro-inflammatory pathways with subsequent release of cytokines and adipokines in cardiometabolic disease and particularly in T2DM is an active area of research interest [32]. Our findings suggest that background polygenic risk for immune-related conditions may heighten risk of incident T2DM development. Of note, we also found that a PRS for T1DM was selected by LASSO models predicting T2DM. A recent study within the Million Veteran Program (MVP) database showed that individuals with T2DM who carry a high genetic risk for T1DM are more likely to have T1DM characteristics including diabetic ketoacidosis, hypoglycemia, and earlier need for insulin therapy [33]. With potential clinical implications for patients who exhibit characteristics of both T1DM and T2DM, these findings underscore a growing need to better understand the shared genetic background between both phenotypes.

Several proteins were repeatedly selected by standard LASSO models across all three traits of interest. In standard LASSO models predicting truncal adiposity and CRF, both FABP4 and LEP carried large beta coefficients, but in opposing effect directions for each trait (positive for truncal adiposity and negative for CRF). FABP4 is mainly expressed by adipocytes and macrophages and has been hypothesized to play a role in lipolysis [34]. Other studies using circulating plasma proteomics have also identified FABP4 as a potential biomarker for related cardiometabolic traits such as IR, obesity, and atherosclerosis [35, 36]. LEP, or leptin, is a known satiety hormone which exhibits increased expression in the setting of obesity and has been associated with IR independent of total body fat mass [37]. Of the proteins selected by standard LASSO models predicting T2DM, GDF15 has previously been associated with a host of diseases within the cardiometabolic spectrum [38]. With known functions in the suppression of food intake and inflammation, GDF15 has become an appealing drug target in the management of obesity, T2DM, and CVD [39].

Aside from incident disease prediction, high-throughput proteomic profiling can further our understanding of the underlying biology of disease and aid in the identification of novel drug targets. Towards the latter aim, we used two-sample MR to infer potential causality of proteins for all three traits of interest. Of proteins with potential causal associations for truncal adiposity, DLL1 has previously been shown to modulate the fate of white and brown adipocytes in mouse models [40]. We also identified LRRC37A2 as potentially causal for CRF. Of note, a variant in the *LRRC37A2* gene was previously identified as a functional candidate in a GWAS of CRF within UKB participants [41]. Finally, in our two-sample MR analysis of T2DM, we corroborated previously known protein-diabetes associations including associations between diabetes with MANSC4, SHBG, and AMY2B [42, 43].

A primary strength of our study is the well-documented stability and reproducibility of the Olink platform [44]. By using a robust clinical risk score such as QDiabetes, we add meaningful findings on whether the incorporation of multi-omics data can enhance existing prediction capabilities for T2DM. Notable limitations of our study include the lack of genetic diversity within the UKB. Additionally, our two-sample MR analyses were restricted to individuals of European ancestry only. Future studies in more diverse populations are necessary to avoid the further perpetuation of health disparities in this line of research. Finally, while others have also shown that plasma proteins can improve cardiometabolic health prediction, we acknowledge potential limitations in using proteins measured in the plasma as opposed to adipose, skeletal muscle, or pancreatic tissues to infer potential causality for our traits of interest [15].

In summary, our findings suggest that plasma proteomic profiling enhances prediction of two risk factors upstream of IR, truncal adiposity and CRF, and provides modest improvements in prediction of T2DM, a downstream consequence of IR. Further large-scale studies in more diverse populations are indicated to better elucidate the advantages multi-omics data could provide for T2DM prediction beyond that offered by the QDiabetes clinical risk score. We also show that the use of substantially smaller proteomic datasets does not significantly compromise prediction performance in comparison to the full proteomic dataset provided by the UKB. Finally, we add a list of potentially causal proteins for truncal adiposity, CRF, and T2DM using expanded proteomic data from the UKB to the existing literature.

## Data Availability

All data produced in the present study are available upon reasonable request to the authors.

## Acknowledgement

The UKB received ethical approval from the Northwest Multicenter Research Ethics Committee and obtained informed consent from all participants at the time of recruitment. This study was conducted under UK Biobank application number 52374.

## Funding and Assistance

This study was supported by a grant from the National Institutes of Health 1R01DK114183. TPG was supported by the Sarnoff Cardiovascular Research Foundation Fellowship.

## Author Contributions

TPG and TLA conceived and designed the study. TPG, ZA, PFK, JZ, and KN carried out the analyses. TPG and TLA drafted the manuscript. TPG, ZA, PFK, JZ, KN, MLC, DJP, RGS, ATH, DS, KW, FA, PST, SLC, and TLA verified the underlying data. TLA is responsible for the integrity of the work as a whole. All authors acquired and interpreted the data, critically revised the paper and had final responsibility for the decision to submit for publication.

## Protein Abbreviations

FABP4: Fatty acid-binding protein, adipocyte
LEP: Leptin
GDF15: Growth/differentiation factor 15
DLL1: Delta-like protein 1
LRRC37A2: Leucine-rich repeat-containing protein 37A2
ABO: Histo-blood group ABO system transferase
PAM: Peptidyl-glycine alpha-amidating monooxygenase
MANSC4: MANSC domain-containing protein 4
SHBG: Sex hormone-binding globulin
AMY2B: Alpha-amylase 2B
TYRO3: Tyrosine-protein kinase receptor TYRO3
NCR3LG1: Natural cytotoxicity triggering receptor 3 ligand 1
TSPAN8: Tetraspanin-8

