## Supplementary Materials for "Plasma proteomic signatures for type 2 diabetes mellitus and related traits in the UK Biobank cohort"

**Electronic supplementary material (ESM)**
**Plasma proteomic signatures for type 2 diabetes mellitus and related traits in the UK Biobank cohort**

### ESM Methods

*Study design and population*

Prevalent type 2 diabetes mellitus (T2DM) was defined as having T2DM at or prior to the baseline visit for those in the fitness and T2DM subcohorts, and at or prior to the imaging visit for those in the adiposity subcohort. Additionally, the QDiabetes score can only be computed in individuals with Townsend deprivation scores (TDSs) between 8 and 14, hemoglobin A1c (HbA1c) values between 15 and 48 mmol/mol, and body mass index (BMI) between $40/2.1^2$ and $180/1.4^2$ $kg/m^2$. Individuals with values outside of these ranges for those variables were excluded from the T2DM subcohort.

*Measurement of clinical variables and polygenic risk scores*

Clinical variables used in the adiposity, fitness, and T2DM subcohorts are listed in the table below. For the QDiabetes clinical score, specifically the QDiabetes-2018 (Model C) function, the following variables were not included: history of gestational diabetes, history of polycystic ovarian syndrome, or history of learning difficulties.

| **Variables** | **UKB field code** |
|---|---|
| *Clinical variables used in the adiposity and fitness subcohorts* | |
| Sex | 31 |
| Age at recruitment | 21022 |
| Self-reported ethnicity | 21000 |
| Body mass index | 21001 |
| Waist circumference | 48 |
| Systolic blood pressure | 4080 |
| Diastolic blood pressure | 4079 |
| LDL cholesterol | 30780 |
| HDL cholesterol | 30760 |
| Total cholesterol | 30690 |
| Triglycerides | 30870 |
| Glycated hemoglobin | 30750 |
| Townsend deprivation index | 22189 |
| Use of blood pressure lowering medication | 6177 |
| Use of cholesterol lowering medication | 6177 |
| Past medical history of hypertension | 2473 |
| Past medical history of cardiovascular disease | 2473 |
| Family history of T2DM | 20107, 20110, 20111 |
| Ever-smoked status | 20160 |
| Self-reported alcohol intake | 1558 |
| Physical activity category | 22040 |

| | |
|---|---|
| *Clinical variables used in the T2DM subcohort* | |
| QDiabetes clinical score | |
| Sex | 31 |
| Age | 21022 |
| Self-reported ethnicity | 21000 |
| Body mass index | 21001 |
| Glycated hemoglobin | 30750 |
| Townsend deprivation index | 22189 |
| Family history of T2DM | 20107, 20110, 20111 |
| Past medical history of hypertension | 2473 |
| Past medical history of cardiovascular disease | 2473 |
| Past medical history of schizophrenia | 130874 |
| Past medical history of bipolar affective disorder | 130892 |
| Past medical history of gestational diabetes mellitus | 4041, 132203 |
| Past medical history of polycystic ovarian syndrome | 130736 |
| Use of a statin medication | 20003 |
| Use of a corticosteroid medication | 20003 |
| Use of a second-generation antipsychotic medication | 20003 |
| Smoking status | 20160, 3456 |
| Systolic blood pressure | 4080 |
| Diastolic blood pressure | 4079 |
| LDL cholesterol | 30780 |
| HDL cholesterol | 30760 |
| Total cholesterol | 30690 |
| Triglycerides | 30870 |
| Self-reported alcohol use | 1558 |
| Physical activity category | 22040 |
| *Standard polygenic risk scores (PRS) used in all three subcohorts* | |
| Standard PRS for age at menopause | 26202 |
| Standard PRS for age-related macular degeneration | 26204 |
| Standard PRS for age at Alzheimer's disease | 26206 |
| Standard PRS for asthma | 26210 |
| Standard PRS for atrial fibrillation | 26212 |
| Standard PRS for bipolar disorder | 26214 |
| Standard PRS for body mass index | 26216 |
| Standard PRS for bowel cancer | 26218 |
| Standard PRS for breast cancer | 26220 |
| Standard PRS for cardiovascular disease | 26223 |
| Standard PRS for celiac disease | 26225 |

| | |
|---|---|
| Standard PRS for coronary artery disease | 26227 |
| Standard PRS for Chron's disease | 26229 |
| Standard PRS for epithelial ovarian cancer | 26232 |
| Standard PRS for estimated bone mineral density t-score | 26234 |
| Standard PRS for glycated hemoglobin | 26238 |
| Standard PRS for height | 26240 |
| Standard PRS for high-density lipoprotein cholesterol | 26242 |
| Standard PRS for hypertension | 26244 |
| Standard PRS for intraocular pressure | 26246 |
| Standard PRS for ischemic stroke | 26248 |
| Standard PRS for low-density lipoprotein cholesterol | 26250 |
| Standard PRS for melanoma | 26252 |
| Standard PRS for multiple sclerosis | 26254 |
| Standard PRS for osteoporosis | 26258 |
| Standard PRS for Parkinson's disease | 26260 |
| Standard PRS for primary open angle glaucoma | 26265 |
| Standard PRS for prostate cancer | 26267 |
| Standard PRS for psoriasis | 26269 |
| Standard PRS for resting heart rate | 21150 |
| Standard PRS for rheumatoid arthritis | 26273 |
| Standard PRS for schizophrenia | 26275 |
| Standard PRS for systemic lupus erythematosus | 26278 |
| Standard PRS for total cholesterol | 21151 |
| Standard PRS for total triglyceride | 21152 |
| Standard PRS for type 1 diabetes | 26283 |
| Standard PRS for type 2 diabetes | 26285 |
| Standard PRS for ulcerative colitis | 26287 |
| Standard PRS for venous thromboembolic disease | 26289 |

Footnote:
a: For self-reported ethnicity, individuals who identified as British, Irish, or any other White background were classified as White; those who identified as African or any other Black background were classified as Black; those who identified as Indian, Pakistani, or Bangladeshi were classified as South Asian; those who identified as Asian or Asian British were identified as Asian; those who identified as having a mixed background were classified as Mixed; all other individuals were classified as Other. b: Body mass index was calculated as whole-body fat mass (kg) divided by the square of standing height (m). c: For both systolic and diastolic blood pressure, two measurements were taken through an automated reading a few moments part. These measurements were averaged together. d: Physical activity was defined as the summed Metabolic Equivalent Task (MET) minutes per week score. Individuals with a MET score of >3,000 minutes per week were classified as having high physical activity, those with a MET

score between 600-3,000 minutes per week were classified as having moderate physical activity, and those with a MET score of <600 minutes per week were classified as having low physical activity. f: Family history of type II diabetes mellitus was recorded if history of diabetes was listed in participants' fathers', mothers', or siblings' list of illnesses. g: Self-reported medication data was used to classify the use of several medications. Statin medication use was recorded if atorvastatin, rosuvastatin, simvastatin, fluvastatin, pravastatin, velastatin, or eptastatin was listed. Corticosteroid medication use was recorded if prednisolone, betamethasone, cortisone, depo-medrone, dexamethasone, deflazacort, efcortesol, hydrocortisone, methylprednisolone, or triamcinolone was listed. Lastly, second-generation antipsychotic medication was recorded if amisulpride, aripiprazole, clozapine, lurasidone, olanzapine, paliperidone, quetiapine, risperidone, sertindole, or zotepine was listed. h: The 36 standard polygenic risk scores (PRSs) listed were generated by the UK Biobank using external genome-wide association study data.

*Measurement of outcomes*

**Adiposity:** Dual energy x-ray absorptiometry (DXA) scans of the whole body, lumbar spine, hip, knee, and lateral spine were performed by trained radiographers using the GE-Lunar iDXA instrument during imaging visits. A quality control assessment of each scan was conducted in real time including checking for image completeness, movement artifact, or presence of foreign bodies. Following this assessment, the acquiring radiographer analyzed each scan and generated numerical measures of bone mass and body composition.

**Fitness:** Fitness tests were performed by trained assessment staff using a stationary bicycle and 4-lead electrocardiogram (ECG) in a random sample of participants during baseline visits. Participants were assigned to 1 of 5 risk categories depending on prior risk factors: Minimal risk, cycle at 50% level; Small risk, cycle at 35% level; Medium risk, cycle at constant level; High risk, take measurement at rest-only; ECG to be avoided, either unsafe or pointless. For participants in categories 1-3, a resting ECG was recorded for 15 seconds during the pre-test stage. In the activity stage, participants were instructed to start pedaling for six minutes while further ECG recording took place. During the recovery stage, participants were instructed to stop pedaling immediately. An additional ECG was recorded during this one-minute period of rest. Fitness tests were terminated if a participant's heart rate reached a pre-set maximum heart level, if participants requested for the test to be stopped, or if participants reported chest pain, dizziness, or feeling faint during the test.

The maximal oxygen consumption ($VO_2max$) estimates used in this study were previously generated by Gonzales et al. using a multilevel modelling framework which incorporated participants' heart rate responses during their fitness tests. The authors developed and validated this cardiorespiratory fitness (CRF) estimation model in a smaller cohort that was age-, sex-, and BMI-matched to the UKB sample undergoing fitness tests. Further details on the methodology and validation of their CRF estimation model can be found in the supplementary materials of their prior publication [1]

*Statistical Analyses*

*Two-sample Mendelian randomization analysis: Cis-protein quantitative trait loci analysis*

We first performed genome-wide association studies (GWASs) for all 2,920 proteins in a cohort of 15,016 UKB participants to identify *cis*-protein quantitative trait loci (*cis*-pQTLs). To avoid potential biases introduced with sample overlap between groups in which the exposure and outcome effect estimates were measured in, this cohort was comprised of participants with proteomics data who had not undergone any tests of interest (including DXA scans and fitness tests) to the laboratory. Prior to GWAS analysis, SNPs with a minor allele frequency (MAF) < 0.001 or an imputation quality score < 0.3 were filtered. Individual NPX values were inverse-rank normalized before GWAS analysis. The GWAS analyses were performed using the REGENIE software, adjusting for several covariates: age, $age^2$, sex, age*sex, $age^2$*sex, proteomic batch, UKB recruitment center, UKB genotype array, the time interval between blood sampling and measurement, and the first 20 genetic principal components [2]. *Cis*-protein quantitative trait loci (*cis*-pQTLs) were defined as SNPs located within 1 Mb on either side of the protein-coding gene. SNPs were pruned based on linkage disequilibrium with a $r^2$ threshold of 0.01. We adopted a significance threshold of $5 \times 10^{-8}$ for defining genetic instruments.

*Two-sample Mendelian randomization analysis: Obtaining effect estimates for each outcome of interest*

To obtain effect estimates for truncal adiposity and $VO_2$max, we performed GWAS analyses of DXA-measured truncal fat percentage and $VO_2$max estimates in a cohort of 33,348 UKB participants and a cohort of 62,402 UKB participants, respectively. Both GWAS analyses were performed with the REGENIE software and adjusted for age, sex, genotype batch, and the first 10 genetic principal components [2]. We inverse-rank normalized both traits prior to analysis. For each GWAS analysis, we filtered SNPs with a MAF < 0.01, minor allele count (MAC) < 100, genotype missingness > 0.1, or a Hardy-Weinberg equilibrium p-value > $10^{-15}$. Samples with more than 10% missingness were also filtered for quality control. Finally, effect estimates for T2DM were obtained from a meta-analysis of summary statistics published by the DIAMANTE (DIAbetes Meta-ANalysis of Trans-Ethnic association studies) consortium ($n_{cases}$ = 55,005 & $n_{controls}$ = 400,308) [3]. These meta-analyzed summary statistics were adjusted for BMI, and limited to results from individuals of European ancestry only. To avoid sample overlap with the UKB group in which *cis*-pQTLs were obtained in, we also opted to use meta-analyzed summary statistics that excluded UKB participants.

*Two-sample Mendelian randomization analysis: Calculating total proportion of variance explained and F-statistics*

We calculated the proportion of phenotypic variance ($q^2$), the total proportion of variance explained ($R^2$), and the *F* statistic using previously established methods described by others[4, 5].

$$q^2 = 2 \times MAF \times (1 - MAF) \times \beta^2$$

where $q^2$ is the proportion of phenotypic variance, MAF is the minor allele frequency, and $\beta$ is the effect size for a given SNP.

$$R^2 = (2 \times MAF \times (1 - MAF) \times \beta^2) / q^2$$

where $R^2$ is the total proportion of variance explained, MAF is the minor allele frequency, $q^2$ is the proportion of phenotypic variance, and $\beta$ is the effect size for a given SNP.

$F = R^2 *(N-1-k) / ((1- R^2) x k)$
where F is the F statistic, $R^2$ is the total proportion of variance explained, N is the sample size, and k is the number of SNPs included in the instrument.

*PRS-BMI Association Analyses*
Interestingly, we found that PRS-BMI was negatively associated with both adiposity and T2DM in LASSO models incorporating clinical variables and PRSs (demonstrated in **ESM Table 1** and **ESM Table 3**, respectively). To investigate these findings further, we first tested individual associations between PRS-BMI with BMI, adiposity, and T2DM. Through regression models, we confirmed that PRS-BMI was positively associated with BMI (β=1.32, SE=0.00676, p-value=0), truncal adiposity (β=0.13274, SE=0.01645, p-value=9.4E-16) and T2DM (β=0.194, SE=0.0234, p-value=9.8E-17). Next, we re-capitulated LASSO models incorporating clinical variables and PRSs and excluded BMI and waist circumference when predicting adiposity and the QDiabetes score when predicting T2DM (as the QDiabetes score includes BMI as a variable) (**ESM Table 7**). In both additional models, we observed that PRS-BMI was selected and had a positive beta coefficient. Ultimately, we hypothesize that the inclusion of collinear variables led to instability in the beta coefficients returned by these models. Through these additional analyses, we show that when collinear variables are excluded, PRS-BMI is associated with both adiposity and T2DM and carries a positive beta coefficient as expected.

**ESM Figure 1. Study populations of each subcohort**

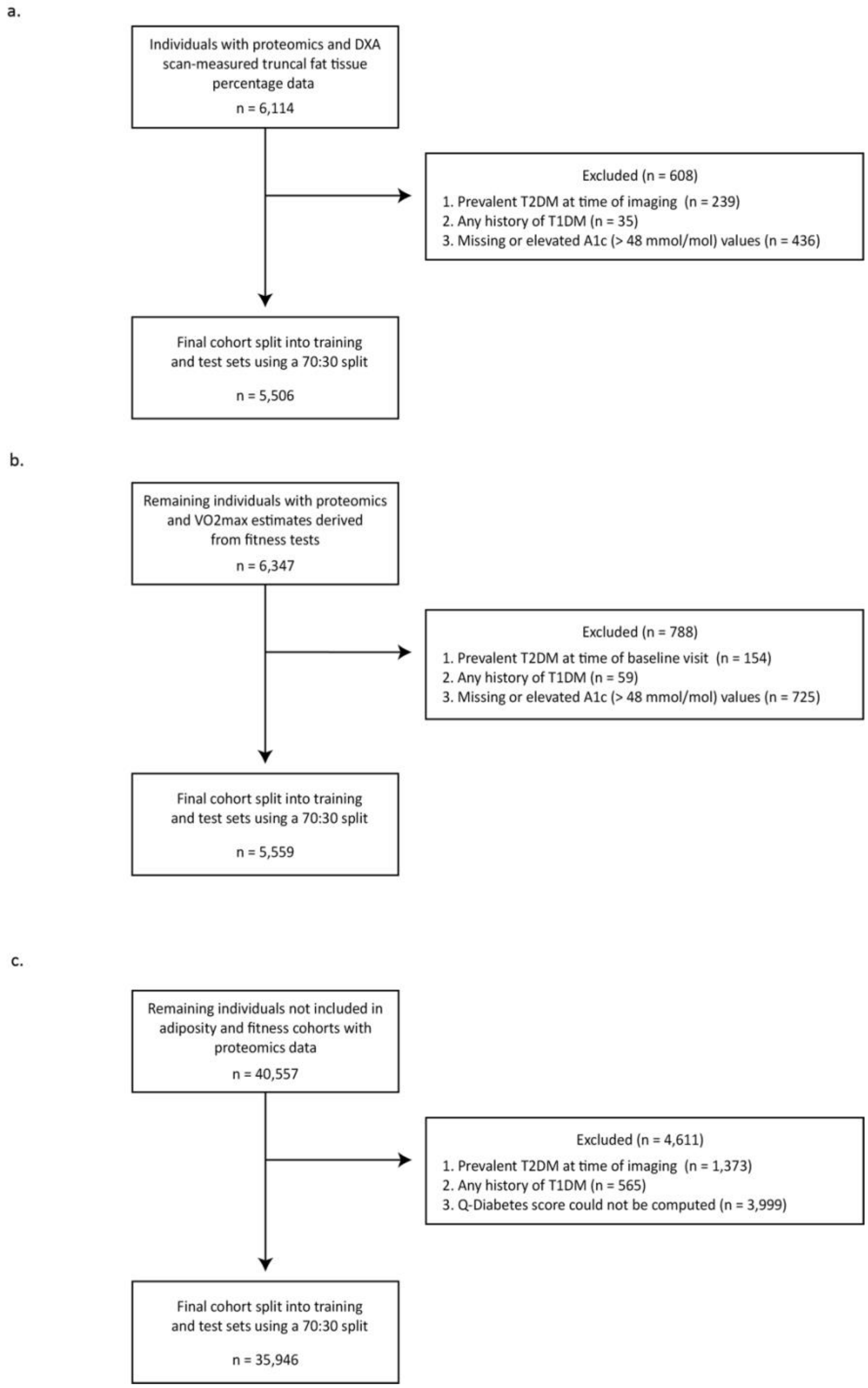


**Footnote:** (**a**) Study population of the adiposity subcohort. (**b**) Study population of the fitness subcohort. (**c**) Study population of the T2DM subcohort. **Abbreviations:** DXA: dual-energy x-ray absorptiometry, T2DM: type II diabetes mellitus, T1DM: type I diabetes mellitus, A1c: hemoglobin A1c, $VO_2max$: maximal oxygen consumption

**ESM Figure 2. Study design for two-sample Mendelian randomization analyses**

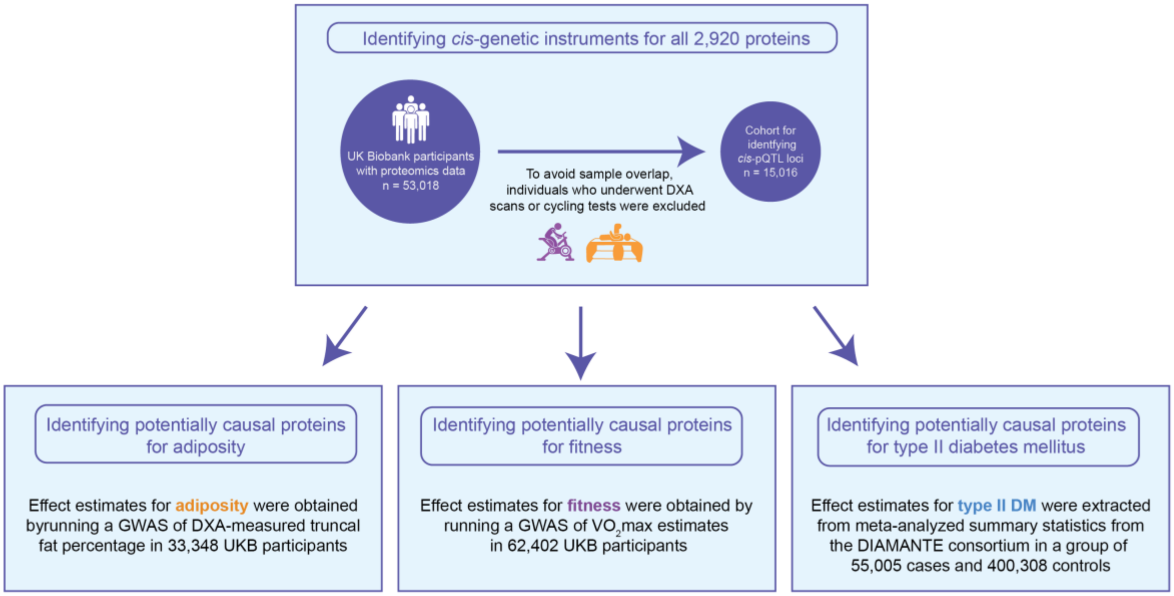


**Abbreviations:** DXA: dual-energy x-ray absorptiometry, *cis*-pQTL loci: *cis*-protein quantitative trait loci, GWAS: genome wide association study, $VO_2$max: maximal oxygen consumption

**ESM Figure 3. Area under the curves (AUCs) of filtered and clustered proteomic datasets in the type II diabetes mellitus subcohort**

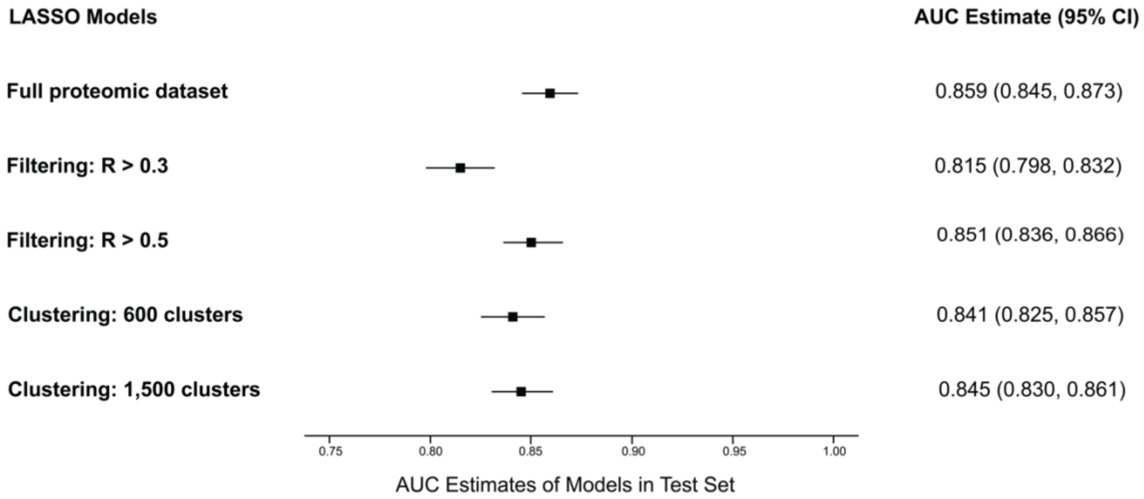


**Footnote:** Models performed in training and test sets of the T2DM subcohort using filtered and clustered versions of the full proteomic dataset. **Abbreviations:** LASSO: least absolute shrinkage and selection operator, AUC: area under the curve

**ESM Figure 4a-d. Manhattan and Q-Q plots for genome-wide association studies of DXA-measured truncal adiposity and cardiorespiratory fitness in UKB participants of European ancestry**

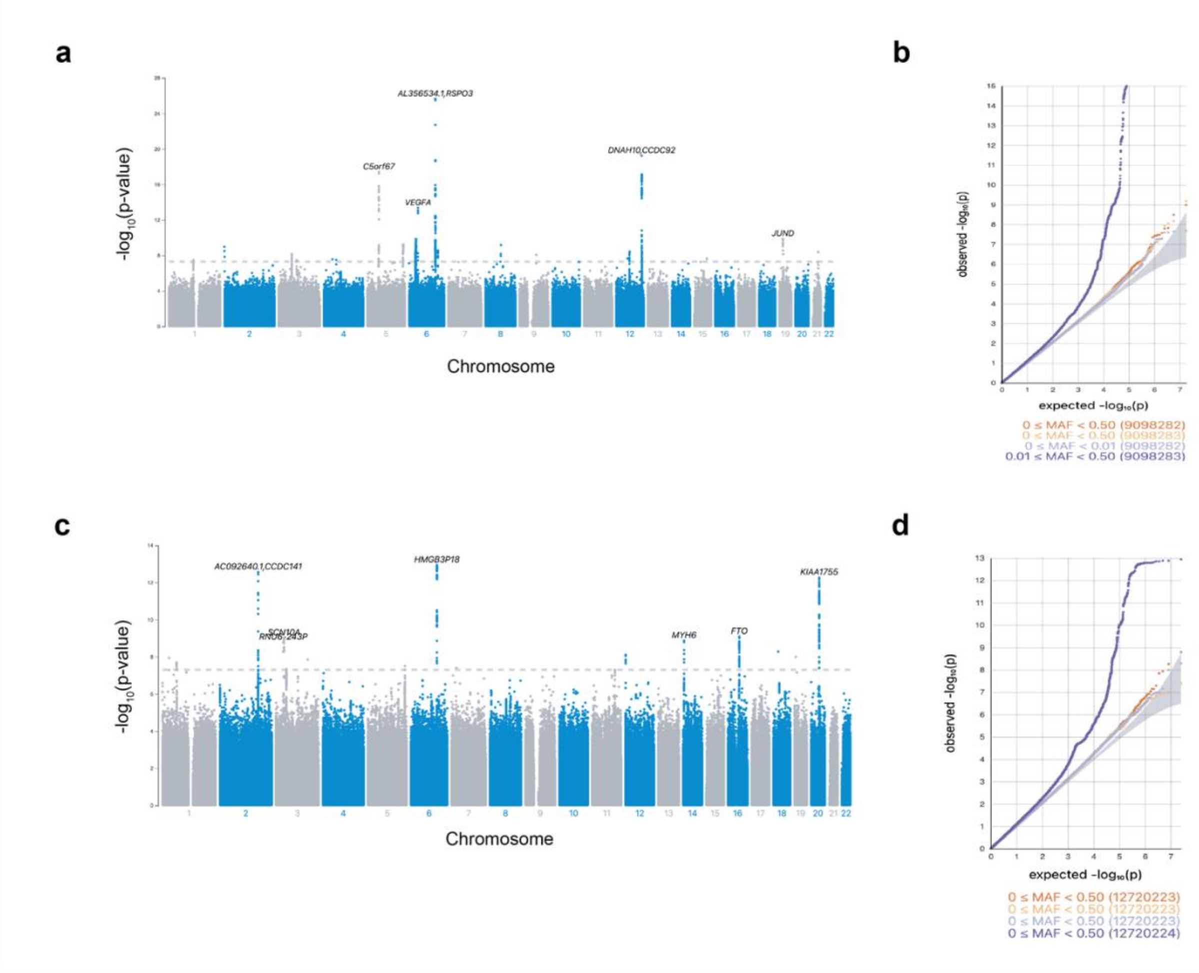


**Footnote:** (**a**) Manhattan plot of GWAS for truncal adiposity. (**b**) Q-Q plot of GWAS for truncal adiposity. (**c**) Manhattan plot of GWAS for cardiorespiratory fitness. (**d**) Q-Q plot of GWAS for cardiorespiratory fitness.

**ESM Figure 5. Cross-validation plots of LASSO regression models in test data sets of the adiposity subcohort with DXA-measured truncal fat tissue percentage as the outcome of interest**

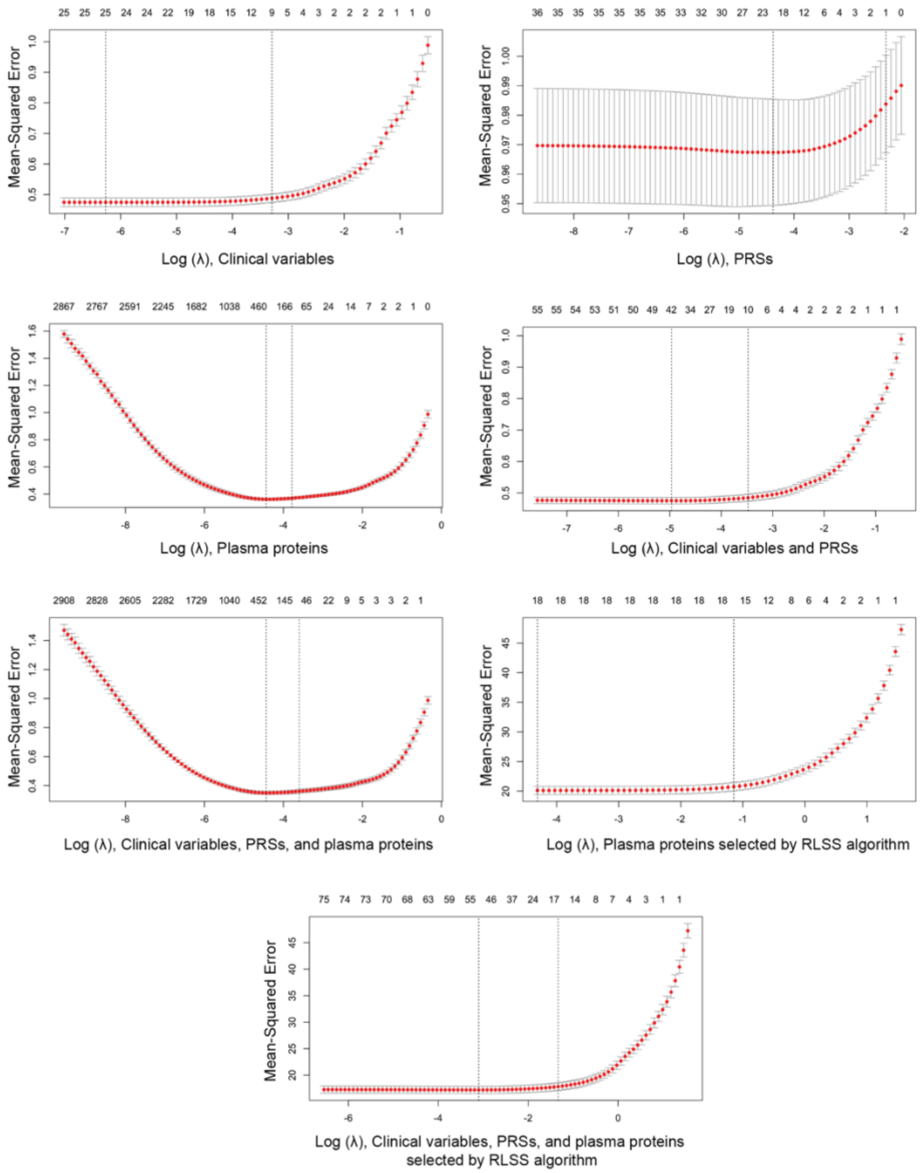


**Footnote:** Upper and lower standard deviation curves along the $\lambda$ sequence (error bars) are shown. **Abbreviations:** PRSs: polygenic risk scores, RLSS: randomized LASSO stability selection algorithm

**ESM Figure 6. Cross-validation plots of LASSO regression models in test data sets of the fitness subcohort with an estimate of $VO_2$max as the outcome of interest**

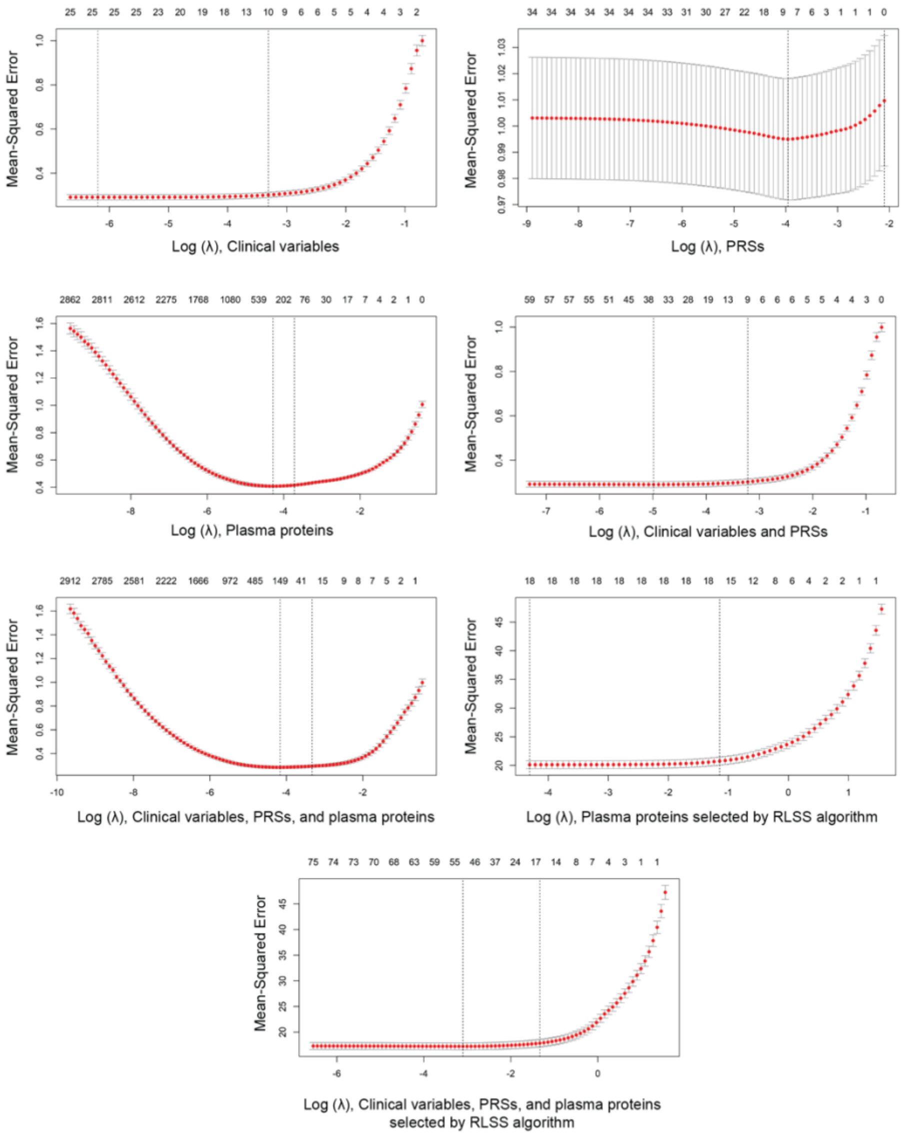


**Footnote:** Upper and lower standard deviation curves along the $\lambda$ sequence (error bars) are shown. **Abbreviations:** PRSs: polygenic risk scores, RLSS: randomized LASSO stability selection algorithm

**ESM Figure 7. Cross-validation plots of LASSO regression models in test data sets of T2DM subcohort with incident T2DM as the outcome of interest**

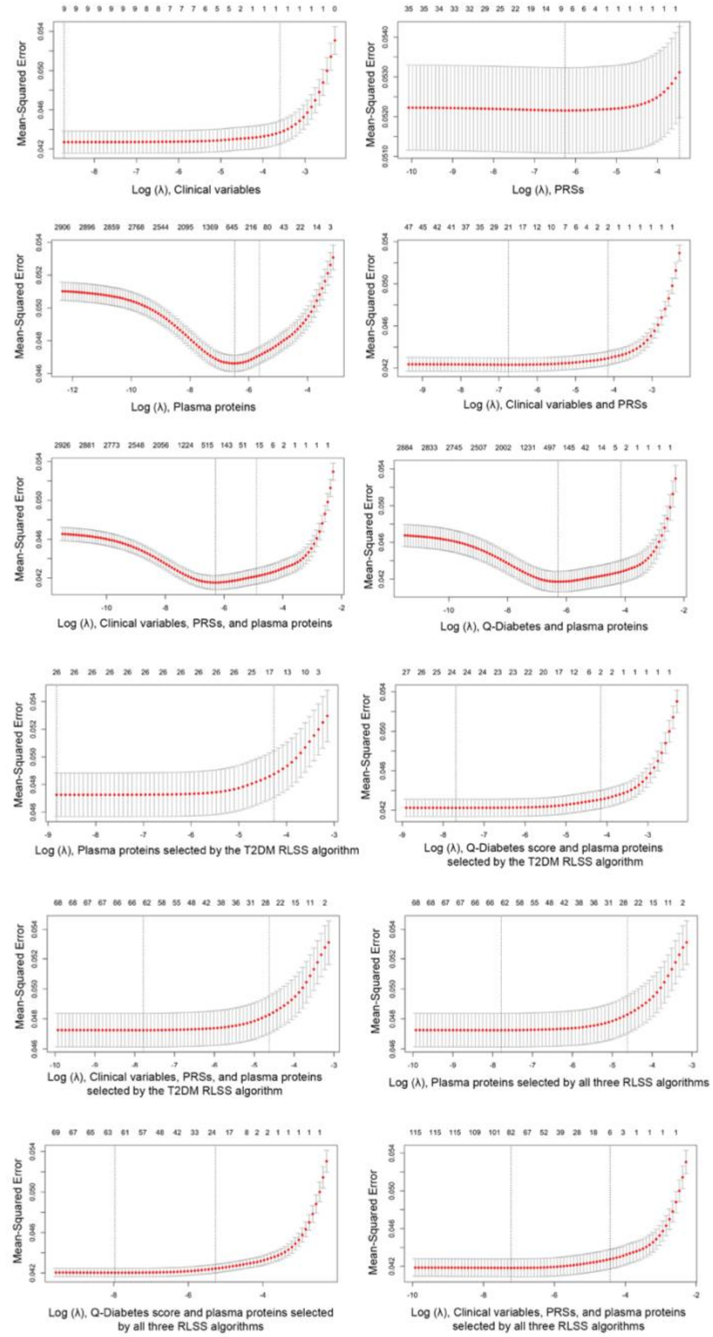


**Footnote:** Upper and lower standard deviation curves along the $\lambda$ sequence (error bars) are shown. **Abbreviations:** PRSs: polygenic risk scores, RLSS: randomized LASSO stability selection algorithm

**ESM Figure 8. Potentially causal proteins for truncal adiposity with results from the inverse variance weighted method and from other sensitivity analyses**

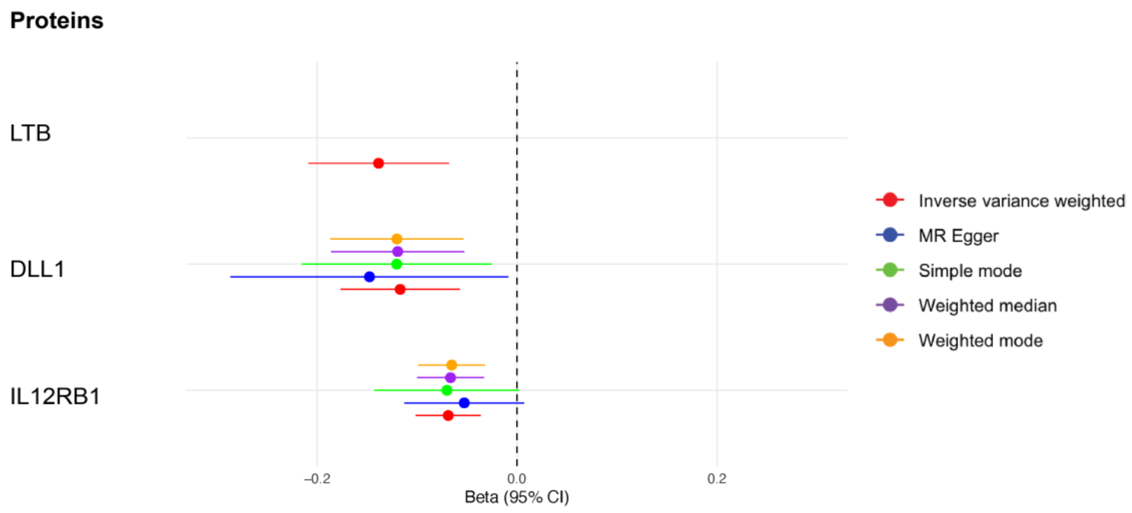


**Footnote:** Forest plot of potentially causal proteins for truncal adiposity based on FDR < 0.5. Red denotes results from the inverse variance weighted method, blue from the MR-Egger method, green from the simple mode method, purple from the weighted median method, and orange from the weighted mode method. **Abbreviations:** 95% CI: 95% confidence interval

**ESM Figure 9. A potentially causal protein for cardiorespiratory fitness with results from the inverse variance weighted method and from other sensitivity analyses**

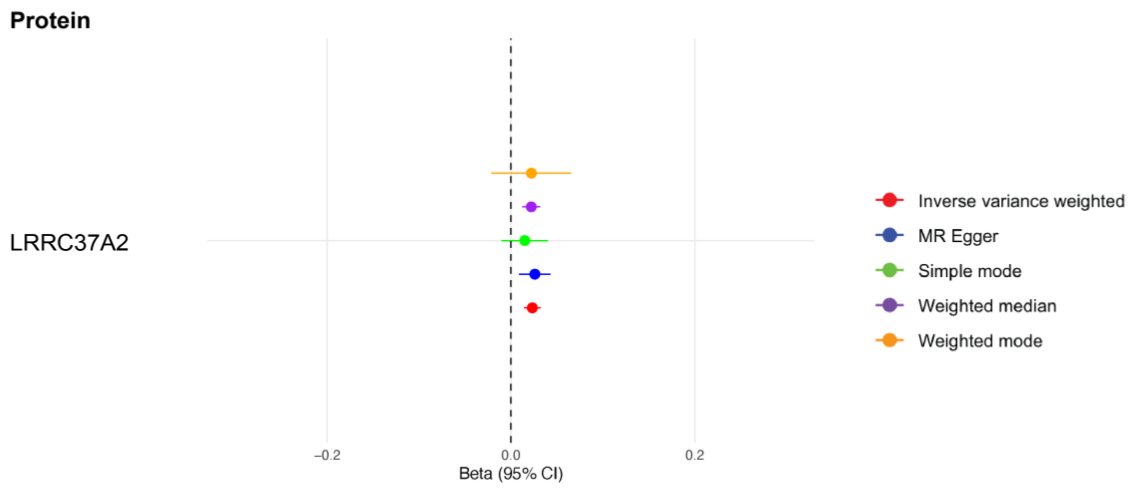


**Footnote:** Forest plot of a potentially causal protein for cardiorespiratory fitness based on FDR < 0.5. Red denotes results from the inverse variance weighted method, blue from the MR-Egger method, green from the simple mode method, purple from the weighted median method, and orange from the weighted mode method. **Abbreviations:** 95% CI: 95% confidence interval

**ESM Figure 10. Potentially causal proteins for type II diabetes mellitus with results from the inverse variance weighted method and from other sensitivity analyses**

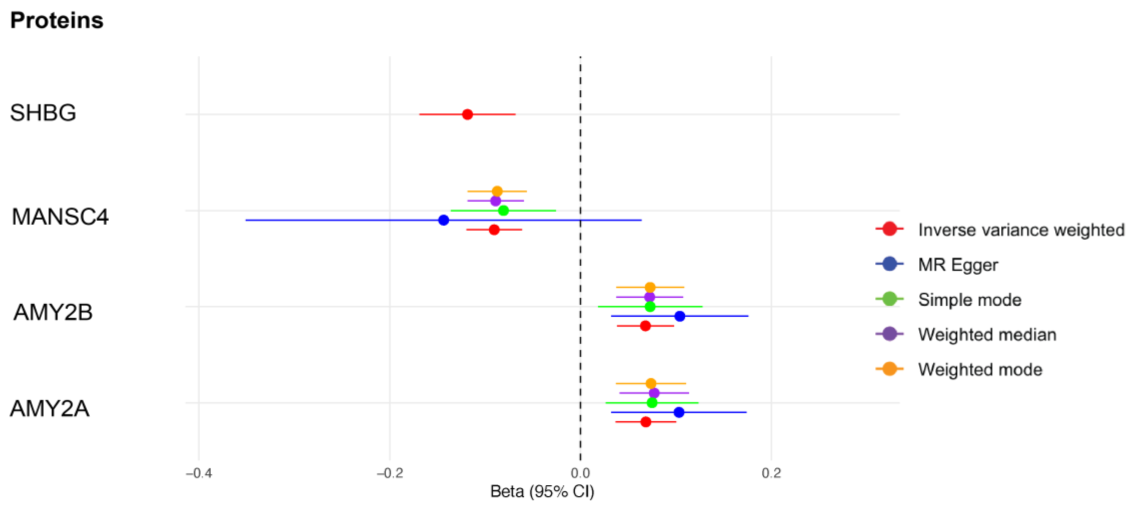


**Footnote:** Forest plot of potentially causal proteins for T2DM based on FDR < 0.5. Red denotes results from the inverse variance weighted method, blue from the MR-Egger method, green from the simple mode method, purple from the weighted median method, and orange from the weighted mode method. **Abbreviations:** 95% CI: 95% confidence interval
